# Childhood adverse experiences associate with reduced brain CB1 endocannabinoid receptor availability in healthy controls and patients with first-episode psychosis

**DOI:** 10.64898/2026.10.26.26366543

**Authors:** Heikki Laurikainen, Lilja Parkkali, Noora Scheinin, Anna Kirjavainen, Olof Solin, Merja Haaparanta-Solin, Raimo K R Salokangas, Jarmo Hietala

## Abstract

**Background:** The brain endocannabinoid system (ECS) is dysregulated in first-episode psychosis (FEP) but understanding of the nature and origins of ECS dysregulation in FEP is limited. The imprinting of developmental adversity and stress into the adult ECS has not been previously studied in humans. We tested the hypothesis that childhood adverse events (ACE) associate with decreased brain type-1 endocannabinoid receptor (CB1R) availability in patients with FEP and healthy controls (HC).

**Methods:** CB1R availability was measured cross-sectionally in male and female patients with FEP (n=15), and healthy participants (n=20). Brain CB1R binding was measured with the selective CB1R radiotracer [^18^F]FMPEP-*d2* using positron emission tomography (PET). Distribution volumes (V_T_) using metabolite corrected arterial input was a proxy of CB1R availability, which was measured in 13 functionally relevant regions of interest (ROI). ACE before the age of 16 were categorized as simple (0-1 events) or multiple (≥ 2 events) using a Finnish population survey questionnaire.

**Results:** An rmANOVA model of regional CB1R V_T_ (ROI ∼ sex + group + sex*group + ACE) indicated statistically significant associations of ACE (p<0.001), group (p=0.027) and sex*group interaction(p=0.018). The largest effect sizes of ACE on CB1R were seen in the amygdala (η^2^=.37), temporal cortex (η^2^=.35), hippocampus (η^2^=.33) and insula (η^2^=.31).

**Discussion:** Multiple childhood adverse effects were associated with lower availability of temporo-limbic CB1R in adulthood in healthy participants and in patients with first-episode psychosis. These results corroborate previous results from pre-clinical studies, and expand the literature in showing that early adversity can be imprinted into the function of human brain in adulthood. The results have implications for studies on resilience and adaptation to stress as well as studies on vulnerability to psychiatric disorders.

## Introduction

Adverse childhood experiences (ACE) are common risk factors for both mental and physical health conditions. It has been estimated that globally about 60% of children are exposed to only 0-1 adverse experiences, and have diminished or unaltered odds of developing a mental health adversity, whereas children with two or more adverse experiences have significantly increased odds for both mental and physical disorders. (Bellis et al., 2019; Felitti et al., 1998; Madigan et al., 2023) ACEs increase the odds of depression symptoms, substance use, social adversity, and they are also associated to a three-fold increased risk for developing a psychotic disorder. (Felitti et al., 1998; Zhou et al., 2025). This has also financial burden consequences. In their systematic review and meta-analysis Bellis et al. (2019) suggested that a reduction of 10% in exposure to ACEs would equate to annual savings of 3 million disability adjusted life years or $105 billion. Individuals with exposure to two or more ACEs seem to account for ∼75% of the costs attributable to ACEs. (Bellis et al., 2019)

The biopsychosocial theoretical framework can be used to understand why ACEs might predispose to morbidity. Psychosocial stress can cause short-term changes and long-term adaptations of biology through a complex interaction with genetic factors. Developmental adversities can thus be embedded into the nervous system as biological diatheses, which can subsequently predispose the individual to psychiatric disorders in the context of homeostatic stress. (Kumsta, 2023) The limbic endocannabinoid system (ECS) has been proposed to act as such a mediator between developmental stress and later biological responsivity to stress. (Lee & Gorzalka, 2015; Lee & Gorzalka, 2012)

The ECS has been proposed to play a key role in stress homeostasis and immune function. (Piazza et al., 2017) The main target of endocannabinoid signals in the nervous system is the type 1 endocannabinoid receptor (CB1R), which is a presynaptic G-protein coupled receptor. The endocannabinoid signalling molecules, such as 2-arachidonoylglycerol and anandamide, are synthesized in the post-synaptic cell secondary to neural transmission, and they act retrogradely on the CB1R to inhibit further neurotransmitter release from the presynaptic neuron. (Lu & Mackie, 2021) The majority of CB1R reside on inhibitory neurons, and to a lesser degree, glutamatergic neurons, whilst some can also be found on other neuron types as well as glial cells. CB1R are ubiquitously expressed throughout the brain, and consistently, the central ECS has been suggested to be a general modulator of behavioural responses to environmental challenges, and regulator of neurodevelopmental processes. (Harkany et al., 2007; Zanettini et al., 2011) However, the ECS has been also implicated in specific functions, such as fear, anxiety, HPA-axis gating, feeding behaviour, cognition, reward and salience. (Kruk-Slomka et al., 2016; Lutz et al., 2015; Mahler et al., 2007; Micale & Drago, 2018)

Animal studies have shown that early-life and chronic stress produces region-specific changes in CB1R with differential effects depending on sex. (Dandi et al., 2024; Lee & Hill, 2013; Reich et al., 2009) Differences of adult ECS phenotype between males and females are plausible also in humans given early evidence on differential CB1R dysregulation in early psychosis. (Laurikainen et al., 2026) Multiple studies show altered peripheral endocannabinoid levels in trauma survivors, (Hauer et al., 2013; Hill et al., 2013) but the complex association between peripheral and central measures in the context of psychiatric disorders complicate interpretations regarding the brain. (Dickens et al., 2020; Hillard, 2017) Human studies on how adult-onset PTSD affects CB1R availability are few, and their results are inconclusive with reports of both unchanged and elevated CB1R availabilities. (Korem et al., 2025; Neumeister et al., 2013) Currently only one pre-print publication by Bassir Nia et al. (2025) reports that exposure to childhood trauma associated with lower limbic CB1R availability in adulthood, measured with [^11^C]OMAR PET. Despite robust epidemiological links between childhood adversity and adult psychopathology, and strong preclinical evidence showing that early adverse events modify brain CB1R signalling, there are no publications directly examining the association of non-traumatic childhood adverse events and in vivo CB1R availability in the adult human brain. Further, although the dysregulation of brain ECS in early or chronic psychosis has been studied in more detail, (Borgan et al., 2019; Ceccarini et al., 2013; Ranganathan et al., 2016; Wong et al., 2010) none have directly examined the role of early adversities in this context despite the associations between developmental adversity, psychosis and the endocannabinoid system.

In this study, we tested whether exposure to multiple ACEs associate with altered brain CB1R availability in a clinical sample of patients with FEP and healthy controls. Our hypotheses were that CB1R availability is lower in individuals with multiple ACEs, and that this effect is more prominent in individuals with FEP. We also explored and controlled for associations between CB1R availability and recent severe adverse life experiences, as well as symptoms of depression.

## Methods

### Participants

All participants were required to be between 18 and 40 years of age. Female participants using oral hormonal contraception were excluded. (Laurikainen et al., 2019) The somatic status of all participants was confirmed by medical examination, blood and urine tests, electrocardiography and MRI. General exclusion criteria for all of the participants were: 1) current Axis I substance dependency disorder, 2) lifetime general medical disorder or injury affecting the brain, 3) illicit substance use within 2 months prior to PET scanning, and 4) pregnancy. Lifetime substance use was documented, and recent drug use was controlled with a urine screen prior to PET. All participants were of Finnish ancestry.

The FEP patients were recruited from psychiatric inpatient wards and outpatient clinics of Turku health services and the Hospital Districts of Southwest Finland and Satakunta. FEP was defined as the presence of DSM-IV Axis I psychosis diagnosis with the onset of treatment for the first episode of symptoms within two years of inclusion. Healthy controls were recruited via the national population registry, a local newspaper advertisement and an advertisement in local educational institutions between 2013 and 2022. Any lifetime psychiatric disorder as according to a structured clinical interview for DSM-IV axis I disorders was used as an additional exclusion criterium for the healthy controls. Altogether 7 male and 8 female FEP patients, and 11 male and 9 female HCs were included in the statistical analyses. Table 1.

**Table 1.**
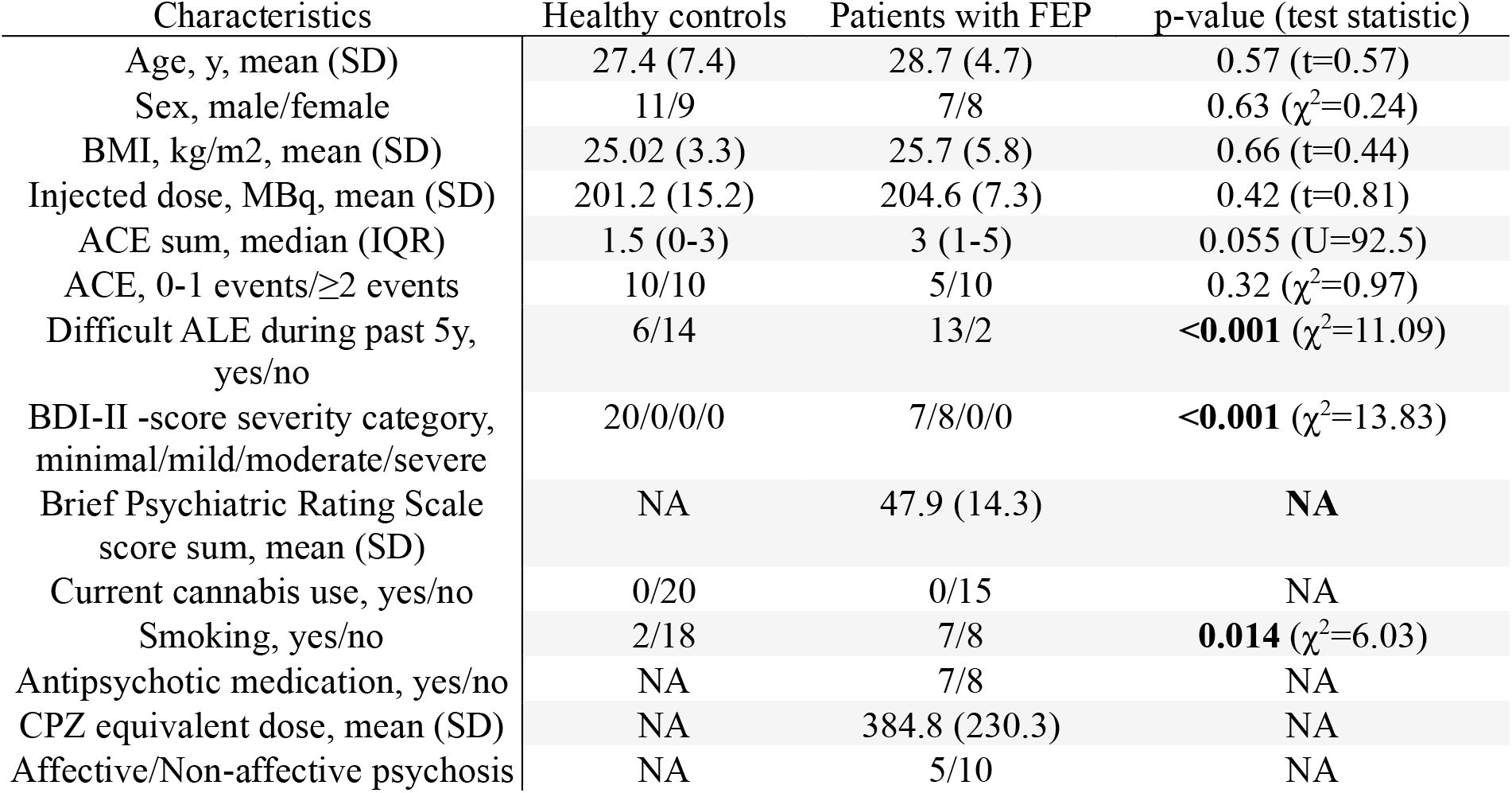
Demographics of the FEP and HC samples.

| Characteristics | Healthy controls | Patients with FEP | p-value (test statistic) |
| --- | --- | --- | --- |
| Age, y, mean (SD) | 27.4 (7.4) | 28.7 (4.7) | 0.57 (t=0.57) |
| Sex, male/female | 11/9 | 7/8 | 0.63 ($\chi^2=0.24$) |
| BMI, kg/m<sup>2</sup>, mean (SD) | 25.02 (3.3) | 25.7 (5.8) | 0.66 (t=0.44) |
| Injected dose, MBq, mean (SD) | 201.2 (15.2) | 204.6 (7.3) | 0.42 (t=0.81) |
| ACE sum, median (IQR) | 1.5 (0-3) | 3 (1-5) | 0.055 (U=92.5) |
| ACE, 0-1 events/$\geq 2$ events | 10/10 | 5/10 | 0.32 ($\chi^2=0.97$) |
| Difficult ALE during past 5y, yes/no | 6/14 | 13/2 | **<0.001** ($\chi^2=11.09$) |
| BDI-II -score severity category, minimal/mild/moderate/severe | 20/0/0/0 | 7/8/0/0 | **<0.001** ($\chi^2=13.83$) |
| Brief Psychiatric Rating Scale score sum, mean (SD) | NA | 47.9 (14.3) | NA |
| Current cannabis use, yes/no | 0/20 | 0/15 | NA |
| Smoking, yes/no | 2/18 | 7/8 | **0.014** ($\chi^2=6.03$) |
| Antipsychotic medication, yes/no | NA | 7/8 | NA |
| CPZ equivalent dose, mean (SD) | NA | 384.8 (230.3) | NA |
| Affective/Non-affective psychosis | NA | 5/10 | NA |

### Imaging measures

CB1R availability was indexed by the distribution volume (V_T_) of the selective radiotracer [^18^F]FMPEP-*d*_*2*_ and PET. Unmetabolized arterial plasma activity used as an input function to model V_T_ using Logan plot. (Logan et al., 1990) [^18^F]FMPEP-*d*_*2*_ was synthesized as described previously with slight modifications. (Donohue et al., 2008) The radiochemical purity was greater than 95% and the molar radioactivity greater than 500 GBq/μmol at the end of synthesis. The MRI and PET procedures, analyses of radiometabolites, imaging and blood data preprocessing, and modelling of V_T_ were done as described previously (Laurikainen et al., 2019) A T1-weighted MRI was acquired using the 3 Tesla Philips Ingenuity PET/MR scanner (TR=8.1ms, TE=3.7ms, flip angle 7°, FOV=256×256×176 mm^3^, voxel size 1×1×1 mm^3^). For three female FEP participants the T1-weighted MRI was acquired using the GE Signa 3 Tesla PET/MR scanner (TR=8.5ms, TE=3.7ms, flip angle 10, FOV=256×256×200 mm^3^, voxel size 1×1×1 mm^3^). Cortical reconstruction and volumetric segmentation were done using the Freesurfer image analysis suite v. 6.0 (http://surfer.nmr.mgh.harvard.edu/). The amygdala (AMY), rostral and caudal anterior cingulate cortex (ACC), frontal cortex (FC), hippocampus (HIPPO), insula (INS), orbitofrontal cortex (OFC), parietal cortex (PAR), posterior cingulate cortex (PCC), prefrontal cortex (PFC), parahippocampus (PHIPP), putamen (PUT), temporal cortex (TEMP) and the thalamus (THA) of the Desikan-Killiany atlas were chosen as the regions of interest.

### Clinical measures

ACEs were assessed by a Finnish population survey questionnaire. (Pirkola et al., 2005) Briefly, the participants were asked to answer “no”, “yes”, or “unable to answer” to 11 questions regarding experiences of adverse events before the age of 16 years; Table 2. All “yes” answers were summed into a variable of lifetime ACE exposure. This sum variable was then dichotomized into categories of: 1) no or singular exposure to adverse events (0-1 ACEs), or 2) exposure to multiple adverse events (≥2 ACEs). The participants also answered a questionnaire concerning lifetime adverse experiences including 19 adverse life events (ALE) with four answer choices: “not at all”, “within the last 6 months”, “within the last 5 years”, or “earlier”. In addition to how recently the participant had experienced the event, they were asked to separately rate the emotional effect of all reported experiences with three answer choices: “not that difficult”, ”difficult”, or “extremely difficult”. An ordinal variable of the number of at least difficult adverse experiences experienced within the last 5 years was computed. Sum scores of the Beck depression inventory (BDI-II) questionnaire were used to measure the severity of depressive symptoms of the subjects at the time of participation in the study. (Beck et al., 1996) Brief psychiatric rating scale (BPRS) symptom scores were calculated by summing individual symptom severity ratings from the 24 item interview. (Nicholson et al., 1995; Sharma et al., 1991)

**Table 2.** Demographics of the ACE groups in a combined FEP and HC sample.

| Characteristics | No or single ACE | Multiple ACE | p-value (test statistic) |
| --- | --- | --- | --- |
| Age, y, mean (SD) | 27.2 (6.6) | 28.5 (6.3) | 0.56 (t=0.58) |
| Sex, male/female | 6/9 | 12/8 | 0.24 ($\chi^2=1.37$) |
| BMI, kg/m<sup>2</sup>, mean (SD) | 24.9 (2.7) | 25.6 (5.5) | 0.63 (t=0.49) |
| Injected dose, MBq, mean (SD) | 204.1 (13.1) | 201.5 (12.1) | 0.54 (t=0.62) |
| Difficult ALE during past 5y, yes/no | 7/8 | 12/8 | 0.43 ($\chi^2=0.61$) |
| BDI-II -score severity category, minimal/mild/moderate/severe | 12/3/0/0 | 15/5/0/0 | 0.73 ($\chi^2=0.12$) |
| Brief Psychiatric Rating Scale score sum, mean (SD) | 48.8 (15.9) | 47.5 (14.2) | 0.88 (t=0.16) |
| Smoking, yes/no | 2/13 | 7/13 | 0.15 ($\chi^2=2.11$) |
| Affective/Non-affective psychosis | 1/4 | 4/6 | 0.44 ($\chi^2=0.60$) |

### Statistical analyses

The normality of regional V_T_ data was assessed using Shapiro-Wilk normality tests. Main effects and within-subject interactions were tested using repeated measures analysis of variance (rmANOVA). Greenhouse-Geisser sphericity corrections were used when applicable. All statistical testing was done using IBM SPSS Statistics v. 31.0.2.0.

The following rmANOVA model was used to test the primary hypothesis:

1: ROI (V_T_) ∼ ACE + sex + group + sex*group

A full factorial model (1), and the following rmANOVA models (1.1-1.2) were used to test the whether the effects of ACE are independent of sex, group, or sex*group interactions, adult adversities and symptoms of depression:

1.1: ROI (V_T_) ∼ ACE + ALE + sex + group + sex*group

1.2: ROI (V_T_) ∼ ACE + BDI-II + sex + group + sex*group

The following rmANOVA model was used to test the sensitivity to known confounders (sex, BMI and age):

2: ROI (V_T_) ∼ ACE + BMI + age + sex

### Voxel-wise modeling and parametric statistical tests

Parametric images of V_T_ were calculated to replicate ROI-level results, and to explore for regional differences of V_T_ not captured by Desikan-Killiany atlas based ROI definitions. The parametric V_T_ maps (1.5×1.5×1.5mm) were generated as detailed in Laurikainen et al. (2019). Briefly, freely available in-house software was used to calculate voxel-wise V_T_ (http://www.turkupetcentre.net/tpcclib/) using Logan plot. The parametric volumes were Gaussian filtered (FWHM 10 mm), and then normalized to standard MNI space. An independent two sample t*-*test was done with SPM 25.01.02 and MATLAB 2026a to explore voxel wise V_T_ differences between the ACE groups without ROI constraints. Three dummy variables were generated as nuisance covariates to adjust for the effects of sex, group and sex*group; contrasts: 1, -1, 0, 0, 0. The uncorrected peak threshold was set at p < .001. The FWER corrected cluster threshold was p < .05, and the extent threshold was adjusted to the size of the smallest significant cluster. To test for sensitivity of the effect for adjusting variables, the parametric t-test was done using only sex or group as nuisance covariates. The probabilistic anatomical assignment of the peak voxels within significant clusters was determined using SPM Anatomy toolbox v. 3.0. (Eickhoff et al., 2005) Overlapping grey matter areas of V_T_ association to ACE and group were explored in a full factorial conjunctional voxel-wise model with sex and group as two-level factors, and ACE as a covariate. Here, the peak threshold was set at p<.001.

### Ethical and other considerations

The authors assert that all procedures contributing to this work comply with the ethical standards of the relevant national and institutional committees on human experimentation and with the Helsinki Declaration of 1975, as revised in 2008. The study was approved by the ethics committee of Southwest Finland (ETMK 98/108/2013), and the Hospital Districts of Southwest Finland and Satakunta. Steps to ensure the confidential handling of data were approved by the ethics committee of Southwest Finland, and the Hospital Districts of Southwest Finland and Satakunta. All participants gave their written informed consent prior to any study procedures. The capacity for consent of patient participants was evaluated by the treating physician and first author. Sex was defined as the sex assigned at birth according to the Finnish national identification number.

## Results

A total of 35 male and female participants were included in the analyses. Except for smoking, there were no differences in key demographics between the clinical groups, or the ACE groups. FEP participants had significantly more difficult adverse life events during the past 5 years, and more severe symptoms of depression, while the ACE groups did not. FEP patients had suffered numerically more adverse childhood events when compared to healthy controls, but the difference was statistically significant only at the trend level. (Table 1 and Table 2)

### The effect of ACE on CB1R availability

A rmANOVA model of regional CB1R V_T_ (ROI ∼ sex + group + sex*group + ACE) showed associations of ACE (F(1,30)=13.69, p=.0009, η^2^=.31), group (F(1,30)=5.42, p=.027, η^2^=.15) and sex*group (F(1,30)=6.26, p=.018, η^2^=.17) and The effect of ACE was regionally differential (ε=.45, F_G-G_(5.37,160.95)=3.24, p=.007, η^2^=.098). The largest effect sizes were seen in the amygdala (η^2^=.37), temporal cortex (η^2^=.35), hippocampus (η^2^=.33) and insula (η^2^=.31).

### Sensitivity analyses

Sex*ACE (p=.66), group*ACE (p=.49) or sex*group*ACE (p=.31) interactions did not affect the significance of ACE (p=0.003), group (p=.019) or sex*group (.013) in a full factorial model. Neither BDI-II depression scale severity (F(1,31)=2.02, p=.16), nor the presence of difficult adverse events during the past 5 years (F(1,31)<.001, p=.99) affected the significance of ACE (p=.002 and p=.002 respectively). Age (p=.70), or BMI (p=.11) did not affect the significance of ACE (p=.002) in model 2.

### Exploration of the effect of singular adverse childhood events on CB1R availability

The “Serious conflicts within the family” item was the only individual ACE to be significantly associated with CB1R V_T_ without adjustments (F(1,33)=13.30, pFWER=0.012, η^2^=.29), and when accounting for sex, group, and sex*group (F(1,30)=14.18, pFWER=0.0078, η^2^=.33). This effect was strongest in the parahippocampal cortex (pFWER=0.004, η^2^=.33), and neither introducing BDI-II depression scale severity, nor the presence of difficult adverse events during the past 5 years affected the significance of the association of this singular adverse event to brain [^18^F]FMPEP-*d*_*2*_ V_T_ (pFWER=0.0086 and pFWER=0.014 respectively).

### Voxel-wise replication and spatial exploration

At a cluster-forming threshold of p<.001 (uncorrected), three clusters (pFWER<.05) showed significantly lower V_T_ in participants with multiple adverse childhood experiences, when compared to the participants with no or singular adverse experiences. The largest cluster (26,314 voxels; 88.8mL; pFWER<.001) extended across the right insula, temporal cortex, parahippocampus hippocampus, amygdala and cerebellum. Two smaller clusters were located in the left fusiform gyrus and cerebellum (5,449 voxels, 18.4mL; pFWER<.001) and temporal cortex and insula (4,948 voxels, 16.7mL; pFWER<.001). Table 3 and Figure 1.

**Table 3.** Significant cluster level results of the effect of ACE on [^18^F]FMPEP-d2 V_T_. The MNI152 coordinates of three peak voxels with the highest T/Z-values, are shown with their respective regional probabilistic location.

| Cluster | k | $p_{FWER}$<br>(cluster) | Region | Side | T | Z | x | y | z |
| --- | --- | --- | --- | --- | --- | --- | --- | --- | --- |
| 1 | 26314 | <.001 | Middle temporal gyrus (46%) | R | 5.05 | 4.26 | 62 | -15 | -28 |
|  |  |  | Inferior temporal gyrus (47%) | R | 4.89 | 4.16 | 51 | -15 | -39 |
|  |  |  | Inferior temporal gyrus (20%) | R | 4.76 | 4.08 | 63 | -38 | -26 |
| 2 | 5449 | <.001 | Temporal occipital fusiform gyrus (70%) | L | 4.28 | 3.75 | -30 | -48 | -20 |
|  |  |  | Cerebellar white matter | L | 4.22 | 3.71 | -20 | -48 | -36 |
|  |  |  | Temporal occipital fusiform gyrus (59%) | L | 4.11 | 3.63 | -28 | -48 | -10 |
| 3 | 4948 | <.001 | Inferior temporal gyrus, posterior division (52%) | L | 4.13 | 3.64 | -46 | -22 | -28 |
|  |  |  | Temporal pole (33%) | L | 4.08 | 3.61 | -44 | 8 | -26 |
|  |  |  | Insular cortex (90%) | L | 3.97 | 3.53 | -39 | -8 | 4 |

**Figure 1.**
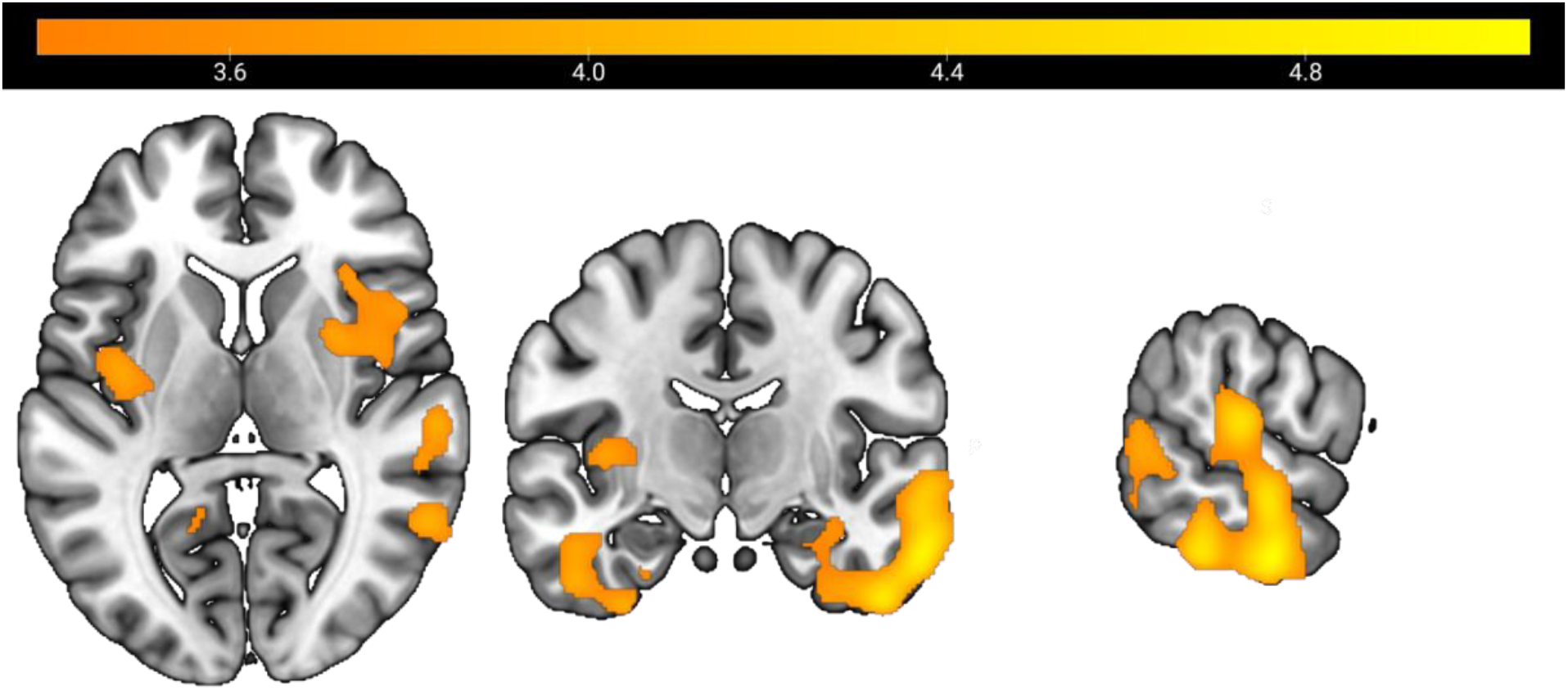
Significant clusters of lower [^18^F]FMPEP-d_2_ V_T_ in participants who reported multiple adverse childhood experiences compared with participants with no or singular adverse experiences. The model was adjusted for sex, group and group*sex as nuisance covariates. The results are shown (x=62, y=-13, z=7) overlaid on a MNI152 T1 template without masking. The color scale and bar shows T-values from threshold at T=3.385 to peak at T=5.05 (p<.001 uncorrected). The displayed clusters are significant at pFWER<.05.

In the exploratory full factor analysis, there were no significant clusters of voxels with conjunct associations of ACE and group to V_T_.

### The association of cumulative exposure to ACE and CB1R availability

The sum of reported ACE was negatively associated to [^18^F]FMPEP-*d*_*2*_ V_T_, adjusted for sex, group and sex*group, in all of the studied ROI using Spearman’s Rho. Although none of the associations survived Bonferroni correction for multiple comparisons, the association remained at trend level in the AMY (ρ=-.46, pFWER=.065). In FEP separately, the sum of reported ACE was negatively associated to [^18^F]FMPEP-*d*_*2*_ V_T_, adjusted for sex, in the ACC, PUT and AMY, but none of the associations survived Bonferroni correction for multiple comparisons. In HC separately, the sum of reported ACE was negatively associated to [^18^F]FMPEP-*d*_*2*_ V_T_, adjusted for sex, in the AMY, OFC, PHIPP and TEMP. None of the associations survived Bonferroni correction for multiple comparisons, but the negative association to AMY remained at trend level (ρ=-.59, pFWER=.078). Supplementary figure 1.

## Discussion

Type 1 cannabinoid receptor availability was significantly lower in participants reporting multiple adverse experiences during childhood. This effect was most prominent in the amygdala-hippocampus-insula-temporal network in both ROI and voxel-wise tests. Notably, the effect of ACE survived after adjusting for sex, group or sex*group. In other words, the effect was present in both healthy controls, and first-episode psychosis patients, with no statistical support for group-specific effects. Further, the effect of ACE was neither sensitive to adjusting for recent symptoms of depression, or exposure to difficult adverse life events. These results point to a persistent, developmentally timed imprint-effect of early adversity on the endocannabinoid system, independent of severe mental disorders.

Our findings are consistent with animal studies showing that developmental stress associates with lasting downregulation of limbic CB1R expression. (Hill et al., 2019; Romano-Lopez et al., 2012) However, our human data did not show evidence of differential effects by sex, as suggested in animal studies such as Suárez et al. (2009). Also, not all animal studies have shown diminished CB1R measures in adulthood, while some have reported increased CB1R mRNA levels. (Robinson et al., 2010) A comprehensive understanding of how different ECS measures relate to each other is difficult to obtain, since receptor availability can dissociate from function depending on cell type, and mRNA, immunoreactivity, protein and availability measures of CB1R seem to provide differential results. (Garani et al., 2021; Steindel et al., 2013)

Our results are consistent with the not yet peer-reviewed preliminary report by Bassir Nia et al. (2025), in which childhood trauma associated with reduced CB1R availability in the hippocampus and amygdala. Our findings extend the previous literature by showing that the adult human ECS is modifiable by developmental non-traumatic adverse experiences. It should be noted that it cannot be ruled out, that genetic differences explain both exposure to adversity and lower CB1R availability, but as a whole the translational evidence base supports a hypothesis on a developmental role of the ECS. Our results also suggest that these developmental effects on the ECS behave predominantly as a trait, which does not invariantly lead to symptoms or disorder in the absence of other predisposing factors. We propose that the temporo-limbic endocannabinoid system is modifiable during the development with long-lasting effects. These results are consistent with the idea that the environment calibrates ECS function during development to facilitate optimal adaptive reactions and behavior. This could explain why the changes in ECS do not invariably associate with a specific disorder, and further suggests that the changes in the ECS might instead be a compensatory effect as discussed by Koethe et al. (2009) in the context of clinical high psychosis-risk states. Previous studies have shown that the brain ECS is altered in first-episode psychosis, and that ACE is a risk factor for the development of psychosis. Based on these notions, we expected a differential effect of ACE on CB1R availability, but this was not the case in our sample. We neither could find evidence suggesting that the cortical areas affected by ACE and early psychosis converge in the same individuals.

It is possible that an altered ECS predisposes to psychiatric symptoms in the presence of other vulnerabilities, extremes of individual allostatic load, or when the adaptive capacity of the ECS is exceeded. Challenges to the ECS in different developmental periods might also lead to differential changes in brain structure and function, since the ECS is involved in multiple neurodevelopmental processes such as regulation of neuronal migration and differentiation, axon pathfinding and synaptic pruning. (Rodrigues et al., 2024; Rubino et al., 2015) Certain developmental periods, which we were not able to distinguish in our ACE data, could thus be more critical for the development of later psychiatric morbidity. Future studies should investigate whether the effects of developmental adversity are temporally and regionally specific for certain developmental periods, and whether this variation translates into differential associations to psychiatric disorders.

### Strengths and limitations

This study was done using validated state-of-the-art methodology for quantifying in vivo CB1R availability in the human brain. The questionnaire for assessing adverse childhood events has been thoroughly characterized and previously implemented in a study of first-episode psychosis patients. (Lindgren et al., 2017) However, the questionnaire is purposed for general population surveys, and as such, it is not an optimal method for the typically modest sample sizes in PET studies. The questionnaire is also not universally adopted as a measure of childhood adversity, which restricts replication attempts, and generalization of the results.

Although the voxel-wise test replicated the ROI-level results in better anatomical detail, the voxel-wise results should be interpreted to indicate the spatial extent of the effect rather than precise anatomical localization according to peak coordinates, given the estimated smoothness of the parametric images (FWHM∼16-22mm).

## Conclusions

This clinical PET imaging study provides early evidence that exposure to multiple childhood adverse events is associated to lower CB1R availability in areas critical for regulation of stress. The association was present in healthy individuals as well as in patients with first-episode psychosis. The association was not explained by recent symptoms of depression or recent difficult adverse life events. Together with emerging findings from other groups, these results tentatively suggest that early adversity could leave a lasting imprint on the limbic endocannabinoid system. The results have clear implications for studies on the neurobiological correlates of resilience and stress adaptation as well as the on studies of environment-biology interaction in psychotic disorders.

## Supporting information

Supplemental results

## Data Availability

All data produced in the present study are available upon reasonable request to the authors.

## Acknowledgements and disclosures

This work was supported by the European Union under the 7th Framework Programme (grant 602478) and the Turku University Central Hospital (grant 11336). We thank the staff of the Turku PET centre, Turku university Hospital, Turku, Finland. All data produced in the present study are available upon reasonable request to the authors.

HL, LP, NS, AK, OS, MH-S, RS and JH designed the experiments. HL, LP, AK and MN performed the experiments. HL, LP, MH-S and OS were involved in experimentation. HL, LP and JH did the data analysis. HL and JH wrote the manuscript. All authors read and approved the submitted version. The authors have no conflicts of interest to disclose.

