## Supplemental results for "Childhood adverse experiences associate with reduced brain CB1 endocannabinoid receptor availability in healthy controls and patients with first-episode psychosis"

| **Supplementary table 1.** Childhood adverse experience questionnaire results | | | |
|---|---|---|---|
| Adverse event | No | Yes | Unable to answer |
| Did your family have long-term financial difficulties? | 23 | 10 | 2 |
| Were your father or mother often unemployed although they wanted to work? | 24 | 7 | 4 |
| Did your father or mother suffer from some serious disease or disability? | 26 | 8 | 1 |
| Did your father have alcohol use-related problems? | 26 | 9 | 0 |
| Did your mother have alcohol use-related problems? | 34 | 1 | 0 |
| Did your father have any mental health problem, e.g. schizophrenia, other psychosis, or depression? | 29 | 4 | 2 |
| Did your mother have any mental health problem, e.g. schizophrenia, other psychosis, or depression? | 28 | 4 | 3 |
| Were there any serious conflicts within your family? | 25 | 9 | 1 |
| Did your parents divorce? | 19 | 16 | 0 |
| Were you yourself seriously or chronically ill? | 34 | 1 | 0 |
| Were you bullied at school? | 19 | 15 | 1 |

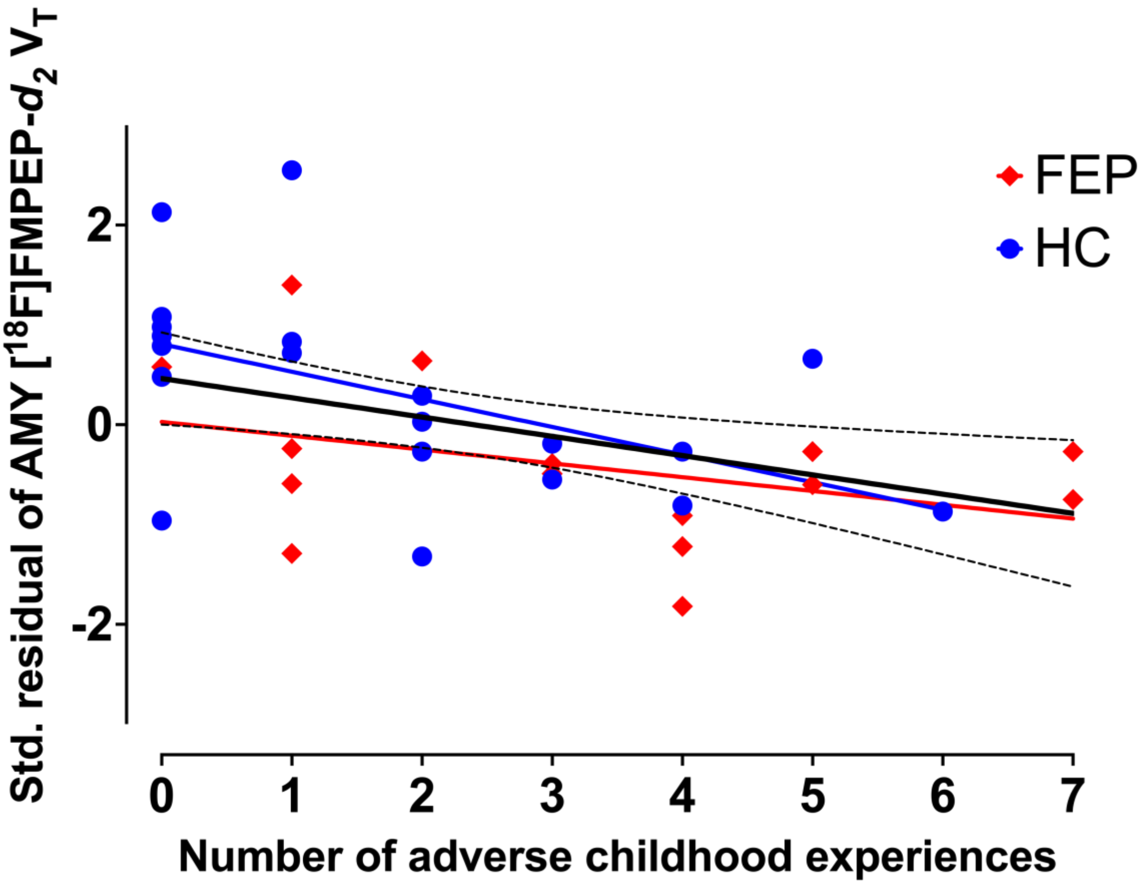


*Supplementary figure 1. The standardized residuals of amygdalar [$^{18}$F]FMPEP-d$_2$ $V_T$ adjusted for sex plotted against the sum of reported adverse childhood experiences. FEP are shown as red diamonds and HC are shown as circles. A linear regression line including both groups is shown as a solid black line with dashed lines indicating 95% CI (Spearman's $\rho$=-.46, $p_{FWER}$=.065; adjusted for sex, group and sex*group). Linear regression lines in red (FEP; Spearman's $\rho$=-.46, $p_{FWER}$=.065), and blue (HC; Spearman's $\rho$=-.59, $p_{FWER}$=.078) show the associations in the groups separately.*
